# Estimating the Daily Milligrams of Morphine Equivalent of Illicit Fentanyl Use in Los Angeles: Clinical and Epidemiological Implications

**DOI:** 10.1101/2025.10.07.25337514

**Authors:** Morgan Godvin, Joseph R. Friedman, Caitlin A. Molina, Adam J. Koncsol, Ruby Romero, David N. Juurlink, Chelsea L. Shover

## Abstract

**Introduction:** The market shift from heroin to illicitly-manufactured-fentanyl in North America led to surging opioid mortality. However, limited information exists about the doses of illicit fentanyl regularly consumed. We examined purity of fentanyl samples and estimate the typical daily oral milligrams of morphine equivalent (MME).

**Methods:** Leveraging community-based drug checking data from Los Angeles, we ascertained the purity of 509 samples of fentanyl collected between September 2023 and January 2026 using liquid chromatography mass spectrometry. We assessed typical consumption quantity and routes of administration among 47 respondents who reported regularly using fentanyl. We estimate bioavailability and MME conversion factors from literature. To estimate daily MME, incorporating all parameter uncertainty, we used a bootstrapping approach with 1,000,000 draws, with sensitivity analyses to assess the impact of factors including the correlation between purity and quantity.

**Results:** Among participants, the mean daily consumption of fentanyl was 1.07 grams (95% prediction interval: 0.03g-4.00g). Illicit fentanyl products had a mean fentanyl purity of 12.47% (0.23%-38.80%), and the mean estimated bioavailability based on routes of administration was 50.82% (30.64%-76.75%). The mean estimated IV fentanyl to PO morphine MME conversion factor was 1 to 183.15 (71.85 - 294.21). The mean estimated daily consumption in our sample was 8,887.55 MME (156.56 MME-41,761.3 MME).

**Conclusions:** Under all plausible estimation scenarios, individuals consuming illicit fentanyl in Los Angeles on average use a quantity of MME several orders of magnitude higher than clinical guidelines or typical methadone doses. This likely contributes to high overdose mortality, high opioid tolerance, and more difficult methadone and buprenorphine induction.

## Introduction

Since the early 2010s, the North American overdose crisis has been driven primarily by illicit fentanyl and its analogues (Friedman and Shover, 2023; Shover et al., 2020). As a synthetic opioid easily produced in large quantities, illicit fentanyl has become a drug of choice for many consumers because it is inexpensive and readily available (Morales et al., 2019). Fentanyl is far more potent than opioids derived from the opium poppy such as heroin and oxycodone, with a milligram of morphine equivalence (MME) conversion factor between intravenous (IV) fentanyl and oral (PO) morphine estimated to range between 1:66 and 1:300 (“Fortnightly Review: Morphine in cancer pain: modes of administration,” 1996; Mercadante et al., 2002; Starlander et al., 2011). It is generally understood that fentanyl’s high potency makes it easy for individuals to inadvertently consume more than intended, leading to greatly elevated overdose death rates(Althoff et al., 2020; Friedman and Shover, 2023). However, detailed information about the actual doses of illicit fentanyl consumed in real-world settings by individuals with opioid use disorder (OUD) is extremely limited. For the use of clinically prescribed pharmaceutical opioids, a daily recommended limit of 90 PO MME is often recommended, although patient specific factors must be considered(Dowell et al., 2016). For substance use disorders involving legal substances like alcohol and tobacco, exposure quantification is part of the standard approach to treatment, with metrics such as drinks per day and pack-years codified in clinical guidelines. In the illicit market, however, exposure quantification is more difficult, given lack of quality control and the shifting composition of illicit drug products (Krotulski et al., 2022). Patients are often unaware of the actual drugs and amounts in their supply, and reliable dosing guidance can be difficult to ascertain. Consequently, there are important potential clinical and epidemiological benefits of quantifying illicit fentanyl consumption among individuals with OUD. These include improved withdrawal management (Thakrar and Kleinman, 2022), improvements in dosing guidance with medications for opioid use disorder (MOUD)(Shearer et al., 2022), and better characterization of the risk environment for this population (Ciccarone, 2017).

Leveraging advancements in community-based drug checking approaches(Delaney et al., 2023; Friedman et al., 2025; Shover et al., 2025a, 2025b) and analytical technologies(Appley et al., 2023; Sisco et al., 2017), we quantified the purity of illicit fentanyl samples in Los Angeles and combined this with self-reported consumption quantities to estimate the daily MME consumed by participants in a drug checking program who reported recent fentanyl use.

## Methods

We analyzed 509 drug samples collected between September 2023 and January 2026 at *Drug Checking Los Angeles*, a community-based drug checking program with multiple sites across Los Angeles County. These samples were drawn from a larger pool of n=2,522 collected during 2023-2026; the subset of 509 represents those for which a) quantitative fentanyl results were available and b) samples were expected by participants to contain fentanyl. Drug samples were provided anonymously and voluntarily by drug checking participants. Services occur in harm reduction settings, and participants provide small quantities of sample for testing. Most samples are expected to reflect the retail drug market, i.e. drugs sold in small quantities to end-users. However, for privacy purposes, we do not inquire if participants participate in drug sales, and may have access to ‘wholesale’ product. For each sample, a few milligrams of drug product were transferred into a vial containing 0.5 mL acetonitrile. Samples were analyzed at the National Institute of Standards and Technology (NIST) using liquid chromatography mass spectrometry (LC-MS) [see previously published methods(Appley et al., 2023; Shover et al., 2025b; Sisco et al., 2017)]. Similar to previous analyses, fentanyl purity was defined as the combined percentage by weight of each sample represented by either fentanyl or fluorofentanyl; these compounds are of comparable potency and impart comparable biologic effects(Canfield et al., 2025).

Drug checking participants were invited to answer a brief, confidential survey about their recent drug use. Among 47 unique respondents who reported using fentanyl in the past 30 days, we assessed consumption (grams per day), and routes of administration (see supplement for more details). For each route of administration, we drew bioavailability estimates from a literature review of experimental data [see supplement](Cook et al., 1993; Darwish et al., 2007; Harris et al., 2003; MacLeod et al., 2012; Mather et al., 1998; Nardi-Hiebl et al., 2021; Streisand et al., 1991; Striebel et al., 1993; Vasisht et al., n.d.). IV fentanyl to PO morphine MME conversion factors were also drawn from literature review [see supplement](Ing et al., 2024; Mercadante et al., 2002; Starlander et al., 2011).

We estimated daily MME consumed by drug checking participants in Los Angeles according to the following formula:

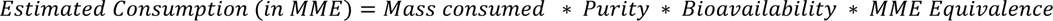

**Equation 1. Calculation of estimated daily milligrams of oral morphine equivalent (MME) used among regular consumers of fentanyl, who visited drug checking services in Los Angeles**

In this equation, 𝑀𝑎𝑠𝑠 𝐶𝑜𝑛𝑠𝑢𝑚𝑒𝑑 is the estimated raw quantity of illicit fentanyl product used per day in milligrams, *Purity* is the proportion of active (i.e. either fentanyl or fluorofentanyl) compound in product expected to be fentanyl, *Bioavailability* is the routes of administration-specific proportion of total drug used estimated to reach the systemic circulation, 𝑀𝑀𝐸 𝐸𝑞𝑢𝑖𝑣𝑎𝑙𝑒𝑛𝑐𝑒 is a metric used to convert doses of different opioids into an approximately equivalent dose of oral morphine to allow for consistent comparisons of potency and risk, which in this case are based on the estimated ratio of physiological potency between IV fentanyl and oral morphine(“Fortnightly Review: Morphine in cancer pain: modes of administration,” 1996; Mercadante et al., 2002; Starlander et al., 2011).

Each of the elements in this equation is associated with a degree of uncertainty. To estimate MME consumed, incorporating uncertainty and individual variation in each parameter, we used a bootstrapping approach. This entailed creating 1,000,000 draws of each parameter—representing the full distribution of uncertainty or variation in each parameter—and calculating MME over these 1,000,000 iterations, yielding a distribution of estimated MME values. At the draw-level, correlation between quantity of fentanyl consumed, and route-of-administration, is preserved from the underlying data, as these quantities are sampled together. When respondents reported employing multiple routes of administration, the fraction of consumption from each route was drawn probabilistically from uniform distributions (which were standardized to sum to 1.0). This assumes that each route of administration was equally likely on average but incorporates uncertainty in the relative proportion of each into the model (see supplement). To generate summary statistics across the draws, we calculate point estimates using means, and 95% prediction intervals (using the 2.5^th^ and 97.5^th^ percentiles) for all estimated quantities. For each, the 95% prediction interval indicates the bounds within which 95% of the estimated parameter for the population fell.

To examine the importance of correlation between purity and quantity of fentanyl consumed, differing estimation scenarios were employed. In a more conservative scenario (sensitivity analysis #1), maximal inverse correlation was induced between quantity of fentanyl consumed and purity of fentanyl consumed, by sorting draws of each quantity in opposite directions (see supplement). This has the effect of assuming that individuals using more potent fentanyl products consume less raw weight than individuals using less potent fentanyl formulation. In a less conservative scenario (sensitivity analysis #2), no correlation was induced, and these two parameters were sampled independently and virtually uncorrelated. This assumes that individuals using more pure fentanyl were no more, and no less, likely to use a greater quantity of fentanyl. Both scenarios are shown as sensitivity analysis in the supplement. In the primary model—displayed in the main text—these two models were combined, by taking 1,000,000 draws from each approach, and calculating summary statistics across the resulting distribution of 2,000,000 draws.

Additional sensitivity analyses were conducted. Sensitivity analysis #3 assumed that for all individuals who employed various routes of administration that included smoking, their estimated bioavailability reflected only values for smoking, as the other routes were negligible. This was conducted as other research has shown a predominance of smoking among individuals employing various routes (Ciccarone et al., 2024). An additional scenario was employed to emulate a behavioral response where consumers attempt to titrate their usage based on potency but do so imperfectly in the fourth sensitivity analysis. In this model, a “noisy” inverse correlation was induced between the daily quantity consumed and fentanyl purity by applying a uniform noise function to the inversely ranked purity distribution (see supplement). Finally, a fifth, “maximally conservative” sensitivity analysis was performed by combining the assumptions of both sensitivity analysis #1 and #3. This model assumed a maximal inverse correlation between quantity and purity alongside a smoking-only bioavailability for those reporting smoking, representing the most restrictive estimation approach.

This project received approval by the UCLA IRB (IRB-22-0760). All analyses were conducted in R version 4.4.1.

## Results

Among the 47 participants reporting past 30-day fentanyl use and consumption quantity, the mean daily consumption of raw fentanyl product (not active ingredient) was 1.07 grams (95% prediction interval: 0.03g-4.00g) (Figure 1). Raw products sold as fentanyl had a mean fentanyl purity (combined concentration of fentanyl and fluorofentanyl, as applicable) of 12.47% (0.23%-38.80%). Almost all participants (n=46; 97.87%) reported smoking/vaporizing fentanyl. Many (n=14; 29.79%) also reported injecting, and 7 (14.89%) reported nasal insufflation. One participant reported exclusively oral ingestion, and one reported oral ingestion, insufflation, and smoking. Assuming probabilistically equal consumption via each method when participants reported multiple routes of administration, and accounting for uncertainty in bioavailability associated with each method (see supplemental methods), the average estimated bioavailability was 50.82% (30.64%-76.75%). The average estimated MME conversion factor between IV fentanyl and PO morphine was 1 to 183.15 (71.85-294.20).

**Figure 1.**
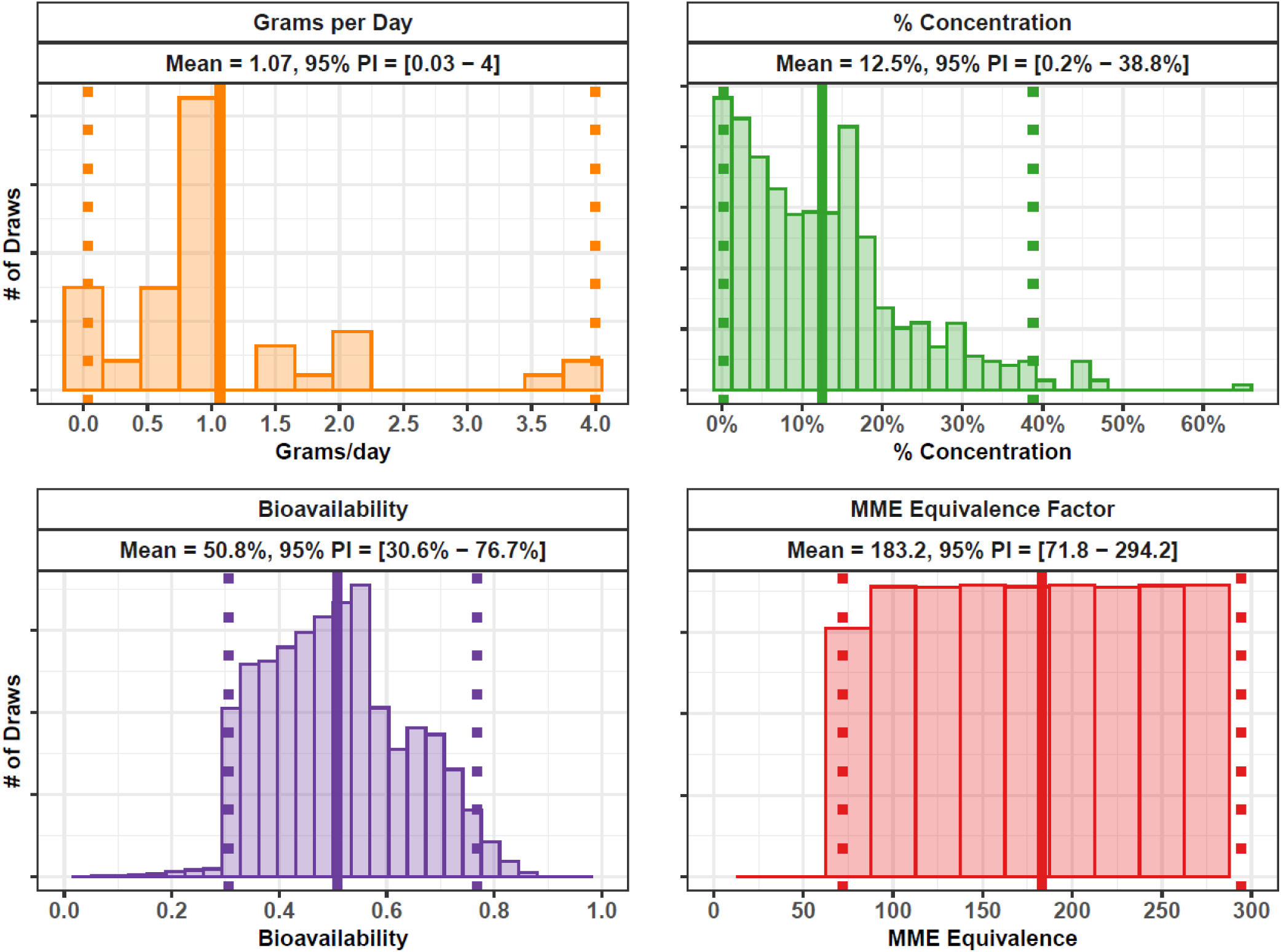
Model Parameters: Fentanyl Consumption, Percent Concentration (Purity), and MME Equivalence. Model parameters are shown via histograms. Each panel represents all 1,000,000 draws from the underlying model. For each parameter, a solid line shows the distribution mean, and dotted lines show the 95% prediction interval (2.5^th^ and 97.5^th^ percentiles). Top Left: The distribution of self-reported grams of illicit fentanyl consumed per day among regular consumers participating in drug checking services in Los Angeles. Top Right: Percent concentration (purity) of expected fentanyl samples provided by clients at drug checking services in Los Angeles. Bottom Left: The estimated bioavailability of participants corresponding to their reported routes of administration (e.g. oral ingestion, smoking, snorting, injecting). Bottom Right: the distribution of MME equivalence factors estimated from literature.

Incorporating each of these parameters, we estimate that participants consumed an average daily estimated MME of 8,887.55 MME (156.56 MME-41,761.30 MME) [Figure 2].

**Figure 2.**
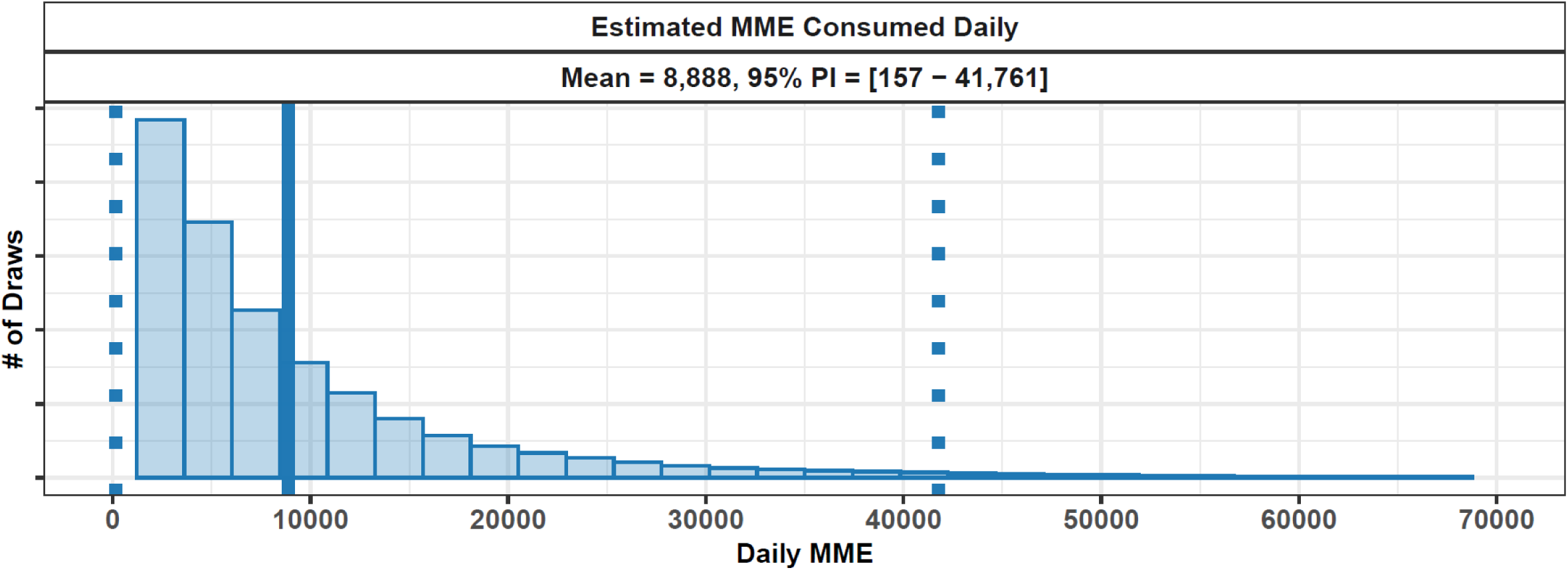
Estimated Daily Fentanyl Consumption in MME. The distribution of the 2,000,000 draws of the model output (estimated daily fentanyl consumption in MME) is shown as a histogram. A vertical solid line shows the distribution mean while dotted lines show the 95% prediction interval (2.5^th^ and 97.5^th^ percentiles).

In the first sensitivity analysis, assuming maximal inverse correlation between quantity and purity, the average estimated MME was lower at 6007.91 MME (537.23 MME - 18,483.31 MME) [Supplemental Figure 2]. In the second sensitivity analysis, assuming no correlation between quantity and purity, the average estimated MME was higher (11,767.19 MME), with a significantly more skewed distribution, reflecting a 95% prediction interval of (59.66 MME - 56,137.06 MME).

In the third sensitivity analysis, assuming smoking was the only route of administration for all individuals reporting smoking and other methods, estimated average MME was moderately lower than the main analysis at 7739.18 MME (130.07 MME - 38,105.49 MME) reflecting lower average bioavailability [Supplemental Figure 3].

In a fourth sensitivity analysis incorporating a ‘noisy inverse correlation’ to simulate imperfect titration behavior (see supplement for methods description and Supplemental Figure 1 for visualization), the estimated average consumption was 7,408.10 MME, with a 95% prediction interval of 316.41 MME to 27,814.71 MME [Supplemental Figure 2].

Finally, in a fifth ‘maximally conservative’ sensitivity analysis combining the assumptions of both Sensitivity Analysis 1 and 3 (see supplement for methods), the estimated average MME was the lowest of our models, yet remained high at 5,125.14 MME (465.36 MME - 14,619.50 MME) [Supplemental Figure 2].

## Discussion

Using drug samples from the real-world illicit drug supply in the Los Angeles area, we found that one gram of a raw product understood to be “fentanyl” in fact contained a range of <1 mg to almost 650 mg of fentanyl. Survey data from people who report recent, regular fentanyl use, report using about 1g of raw product use per day on average, consistent with previous estimates (Ciccarone et al., 2024). Combining these parameters, we estimate that a typical consumer of illicit fentanyl uses the equivalent of almost 9,000 milligrams of oral morphine daily, an extraordinary amount when considered in light of treatment guidelines that discourage doses in excess of 90 milligrams daily for the treatment of chronic pain (Dowell et al., 2016).

It is generally recognized that as a result of tolerance, opioids have no set upper dose threshold, and that usage quantities can escalate rapidly among individuals with OUD who have ample access to opioids. It has also been previously described that the illicit market shift from heroin to illicit fentanyl drastically increased the potency of illicit opioid products, a key factor in skyrocketing rates of overdose and mortality (Ciccarone, 2017; Friedman and Shover, 2023). To our knowledge, this analysis represents the first effort to quantify these phenomena in estimated MME consumed per day among individuals consuming illicit opioids in the fentanyl era.

The advent of community-based drug checking techniques provides a powerful opportunity to open the ‘black box’ of illicit drug products, and quantify drug contents with accuracy and precision (Bailey et al., 2023; Delaney et al., 2023). We find that the typical product sold in Los Angeles as fentanyl, and expected to be illicit fentanyl, is about 12.5% pure. One gram of this product, sold for approximately $100 USD in Los Angeles, would contain about 125 mg of active fentanyl, roughly 100-fold higher than a typical daily amount used for IV analgesia in an opioid-naïve adult. However, purity of illicit fentanyl samples in Los Angeles is also highly variable, ranging from 0.1% to almost 65% among quantified samples. A gram of illicit fentanyl could therefore contain from less than 180 MME to almost 120,000 MME. Assumptions about bioavailability, MME conversion factors, and the correlation between parameters, do influence the overall estimated dosage. However, under all plausible scenarios, individuals consuming illicit fentanyl in Los Angeles on average appear to be consuming MME ranges many orders of magnitude above clinical dosing regimens.

This high—and highly variable—potency illustrates the impact of illicit fentanyl on the risk environment for people with OUD. The extraordinarily elevated overdose risk in the fentanyl era can be understood, in part, as a logical consequence of extreme variations in purity, wherein a gram of street product can vary wildly in its overall opioid content.

Our findings illustrate how insights gleaned from drug checking data can have potential implications for clinical practice—in particular, withdrawal management and stabilization of patients initiating MOUD. High average MME content of illicit fentanyl samples is reflected in the extremely elevated tolerance noted among people using illicit opioids. Many individuals report difficulty returning to heroin use after initiating illicit fentanyl use (Friedman et al., 2022a). Similarly, MOUD dosing requirements have also increased sharply in the fentanyl era (Bolshakova et al., 2024; Shearer et al., 2022; Tsui et al., 2025). Of note, these results do not imply that individuals stabilized on methadone require doses that fully equal the MME of the illicit fentanyl that they had been consuming; due to cross-tolerance when switching opioids, as well as differences between the steady-state induced by methadone and the pulsatile plasma concentrations induced by illicit fentanyl use, sufficient methadone doses may only represent a much smaller fraction of MME. Nonetheless, quantifying the MME of illicit fentanyl consumed by patients may be predictive of the ultimate dose of methadone needed.

Methadone and buprenorphine are the best evidence-based treatments for OUD, and their use is associated with marked reductions in mortality rates (Santo et al., 2021). Nevertheless, a large and growing body of evidence suggests that MOUD initiation has become more difficult in the fentanyl era, with uptake of MOUD and engagement in treatment among people with OUD remaining low in the United States (Jones et al., 2023). Our results suggest that a major reason for this may be the relatively lower MME provided by typical methadone doses compared to typical illicit fentanyl consumption patterns. For instance, typical starting doses of methadone for OUD, ranging from 20 to 40mg daily, are approximately equivalent to 94-188 MME daily. Even a high-end dose of 180mg of methadone daily would be equivalent to 846 MME, representing only a small fraction of the estimated almost 9,000 MME consumed daily by a typical participant in our sample. Methadone doses in this range are unusual, in part because of concerns about dose-dependent QT prolongation.

Although it is generally not necessary or clinically appropriate to fully replace illicit opioid dosing during MOUD initiation, the provision of higher induction and maintenance doses of methadone, equivalent to a larger fraction of MME consumed via illicit fentanyl ingestion, may be warranted among patients using illicit fentanyl. Alternatively, augmenting methadone with other agents, such as slow-release oral morphine, may have similar benefits without the QT prolongation risk (Klimas et al., 2019). This may also help explain improved outcomes seen by some providers when initiating buprenorphine with higher doses, i.e. ‘macrodosing’ (Tsui et al., 2025), although the unique pharmacological properties of buprenorphine make a direct MME comparison more difficult. Through a nuanced understanding of their patients’ illicit fentanyl consumption patterns, and local drug checking results, physicians prescribing MOUD initiation may be able to better tailor dosing to improve patient comfort and improve retention in treatment.

### Limitations

The illicit fentanyl risk environment in Los Angeles is similar to many other locations on the West Coast—characterized by a later arrival of illicit fentanyl compared to most of the country (Shover et al., 2020), a delayed albeit large-magnitude increase in overdose deaths after the advent of illicit fentanyls (Friedman and Shover, 2023), a more recent arrival of xylazine as a co-adulterant (Friedman et al., 2022b, 2025), and a predominance of smoking over injection use (Ciccarone et al., 2024; Eger et al., 2024). Although the illicit drug market in LA is broadly reflective of trends in the West Coast, the data leveraged here may have certain biases. Information collected from community-based drug checking programs is subject to convenience sampling and may not be representative of the illicit drug supply in Los Angeles as a whole. The purity of fentanyl likely varies by geography. Consumption patterns may be higher among drug checking participants compared to other people who use opioids, as this likely represents a population with high-volume illicit fentanyl intake. Additionally, we only included fentanyl potency from two fentanyl analogs (fentanyl and fluorofentanyl), which composed the vast majority of our sample, and which were assumed to be equipotent. Literature suggests that this is a generally reasonable assumption (Canfield et al., 2025), however it may have had a small effect on the study results. For simplicity we did not include a small number of other fentanyl analogs contained in our samples (e.g. 14 quantified carfentanil samples, mostly at very low concentrations) (Shover et al., 2025b). This may have slightly underestimated the true overall potency of illicit fentanyl samples in our study population.

We used literature sources to describe a range of plausible values for bioavailability and MME conversion factors. These factors considerably affect study results and yet limited high-quality data are available to guide their selection. Where possible, we used conservative estimates for these parameters.

Sensitivity analyses highlight that, like most simulation or bootstrapping exercises, our results do change when parameter assumptions are modified. Nevertheless, under all plausible parameter scenarios, the estimated MME of fentanyl consumed by this population remains extremely high. However, refining parameter assumptions will be an important area of future work.

When comparing these estimates to the clinical guideline of 90 MME daily, it is notable that the lower bounds of the 95% prediction intervals vary significantly depending on model assumptions. In the scenario assuming no correlation between quantity and purity (Sensitivity Analysis 2), the 95% prediction interval encompasses the 90 MME clinical threshold. In contrast, models incorporating an inverse correlation between quantity and purity produce lower bounds that consistently remain above this guideline. This nuance highlights that while the average estimated consumption is staggering, the extreme variability of the illicit supply means that a subset of consumers may be exposed to daily MME totals much closer to clinical maximums, and clinicians should not assume that all individuals that use fentanyl have tolerance levels that are extremely elevated.

One very impactful decision relates to the degree of correlation between fentanyl quantity and purity consumed. In the supplement we present sensitivity analyses, assuming various scenarios. The most extreme include: 1) strong inverse correlation between these parameters (i.e. people using stronger product use less), and 2) no correlation. Given the lack of quality data to differentiate between how correct these scenarios are, we prefer to include both in the final model, combining the distributions to generate final estimates. This approach recognizes that uncertainty from both of these scenarios is worth including in the model, and the true degree of correlation is likely somewhere between these two. Although we suspect the no correlation scenario is likely more accurate, we prefer to present the more conservative hybrid approach in the main text. The lack of a large sample of linked purity and quantity values represents an important limitation of this study. Larger samples may be able to directly measure this correlation in future studies. Additionally, more data about how these quantities vary by participant characteristics, e.g. race/ethnicity, gender, etc. represent important areas of future study. Bioavailability parameters are also consequential for the results. However, given the overall extreme magnitude of estimated MME in our results, these assumptions are unlikely to change the main study conclusions. Further research is needed to better characterize the bioavailability and other pharmacodynamic and pharmacokinetic properties of illicit fentanyl vaporized or smoked via different methods (e.g., pipe, foil, dabbing equipment) via updated laboratory methods. More data is also needed to characterize fentanyl usage patterns throughout the day, e.g. frequency of dosing, and how this affects total MME consumed, tolerance, and MOUD initiation. Additionally, the role of underlying disease processes (e.g., COPD, asthma, pulmonary hypertension) on illicit fentanyl dosing and absorption deserves consideration.

### Conclusions

We provide the first effort, to our knowledge, to quantify the MME consumed by people who regularly use illicit fentanyl. We find that the average consumption of almost 9,000 MME is vastly higher than clinical guidelines (which often recommend limiting opioid use to 90 MME) or typical methadone doses. This may explain highly elevated overdose mortality rates in the fentanyl era, as well as increased difficulty of MOUD initiation. Further research is needed to assess the degree to which these findings generalize beyond this high acuity population in Los Angeles.

## Funding

CLS received support from the National Institute on Drug Abuse (K01DA050771). This work was supported by the Centers for Disease Control and Prevention as part of Overdose Data to Action: LOCAL (CDC-RFA-CE-23-0003), and made possible through an equipment grant from the James B. Pendleton Charitable Trust to the UCLA AIDS Institute and UCLA Center for AIDS Research. JRF received funding from the National Institute on Drug Abuse (1U01DA063078). AJK received educational support through the NIH/National Center for Advancing Translational Science (NCATS) UCLA CTSI (TL1TR001883). The funders played no role in the design and conduct of the study; collection, management, analysis, and interpretation of the data; preparation, review, or approval of the manuscript; and decision to submit the manuscript for publication. CLS and JRF had full access to all the data in the study and take responsibility for the integrity of the data and the accuracy of the data analysis.

## Conflicts of Interest

Authors declare no conflicts of interest.

## Data Availability

Individual-level data in this study are sensitive and may not be shared. Summary statistics may be requested upon reasonable request to the authors.

## Supplement

### Supplemental Methods

#### Purity

1. Data describing fentanyl purity and daily quantity ingested among regular consumers were assessed via data generated from anonymous participants accessing a community-based drug checking program, *Drug Checking Los Angeles*. Participants voluntarily provided samples of illicit drug products for testing at multiple different sites in Los Angeles County, California from September 2023 to January 2026.
2. Samples were analyzed initially in the field with Fourier-transform infrared (FTIR) spectroscopy and immunoassay test strips. Samples were then sent to the National Institute of Standards and Technology (NIST) for secondary laboratory-based qualitative and quantitative testing using direct analysis in real time mass spectrometry (DART-MS) and liquid-chromatography mass spectrometry (LC-MS). Both FTIR and DART-MS assess samples against libraries of over 1,300 substances, including pharmaceutical and illicit drugs, adulterants, cutting and bulking agents, precursor chemicals, and other substances (e.g., adhesives, food products, etc.). The LC-MS quantification panel initially included twelve substances: fentanyl and fluorofentanyl, fentanyl precursor chemicals (4-ANPP, phenethyl 4-ANPP), heroin, methamphetamine, cocaine, α2- agonists (xylazine and medetomidine), and three common fentanyl adulterants (tetracaine, lidocaine, and Bis(2,2,6,6-tetramethyl-4-piperidyl) sebacate (BTMPS), and was later broadened to include additional fentanyl analogs and other substances of interest in December 2024. Lidocaine and BTMPS were added to the quantitation panel in July 2024.
3. Of n=2,522 total available samples, this study included n=509 that were sold as fentanyl, expected to contain fentanyl, per client self-report, and had quantified results available based on LC-MS for fentanyl, fluorofentanyl, or both. Only n=4 samples contained fluorofentanyl but not fentanyl.
4. Samples which were designated to be below the LC-MS limit of quantitation were imputed to be 0.1% by mass for purposes of analysis.
5. The 509 available fentanyl purity values were resampled with replacement to create 1,000,000 draws representing the distribution of purity using the sample() function in R. A seed was set to ensure the reproducibility of results between code iterations.

#### Drug Quantity Consumed

1. Information on drug quantity consumed for fentanyl (and other drugs) were collected through an anonymous, optional survey conducted by trained drug checking staff.
2. A total of n=47 participants who regularly consume fentanyl self-reported the quantity of fentanyl product they consume in grams or dollars, and had the option to report the quantity per day, week, or month. Weekly values were converted to days by dividing by 7. Monthly values were converted to daily by dividing by 30. Responses given in dollars were converted to grams using a standardized price of $100 USD per gram (which is somewhat cautious, and may underestimate the quantities that participants obtain per dollar). They also self-reported which of the following routes of administration they use to consume fentanyl: inject (IV or IM), smoke, snort, oral, or rectal.
3. The 47 available fentanyl quantity values (alongside associated route of administration information from the same individuals) were resampled to create 1,000,000 draws representing the distribution of daily fentanyl consumption using the sample() function in R. In this fashion route of administration and quantity consumed are considered in a linked fashion, not sampled independently.

#### Bioavailability and Equivalence

1. Literature values were used to estimate distributions for all other model parameters, including bioavailability and MME conversion factors.
2. Bioavailability of orally consumed fentanyl was estimated using the below studies. Each use distinct formulations of fentanyl designed for buccal absorption. They may not be fully comparable to powder fentanyl absorbed via buccal mucosa and the GI tract. Overall, the oral bioavailability was estimated using a normal distribution with a mean of 30%, with a standard deviation of 10%, which is larger variance than the below studies, but accounts for uncertainty due to differences in formulation.

a. Vasisht, N., Gever, L.N., Tagarro, I., Finn, A.L., n.d. Single-Dose Pharmacokinetics of Fentanyl Buccal Soluble Filmpme_8751017..1023.

i. This study estimated oral bioavailability at 35%.
b. Darwish, M., Kirby, M., Robertson, P., Tracewell, W., Jiang, J.G., 2007. Absolute and Relative Bioavailability of Fentanyl Buccal Tablet and Oral Transmucosal Fentanyl Citrate. The Journal of Clinical Pharma 47, 343–350. https://doi.org/10.1177/0091270006297749.

i. This study estimated oral bioavailability of a formulation of oral fentanyl at 31%, with a standard error of 3.6%.
c. Streisand, J.B., Varvel, J.R., Stanski, D.R., Maire, L.L., Ashburn, M.A., Hague, B.I., Tarver, S.D., Stanley, T.H., 1991. Absorption and Bioavailability of Oral Transmucosal Fentanyl Citrate. Anesthesiology 75.

i. The oral bioavailability of fentanyl was estimated at about 33% absorbed through the gastrointestinal tract.
3. Bioavailability of vaporized (often referred to colloquially as ‘smoked’) fentanyl was estimated from the below studies (A and B), which ranged from 78%-100% bioavailability when using specialized devices to vaporize fentanyl at a precise temperature and minimize vapor lost to the environment. Therefore, these studies would represent a reasonable upper bound of bioavailability, likely more efficient than real-world conditions. It was therefore additionally assumed that an additional quantity of the total drug is left in the pipe/on the foil, or lost to the environment (based on evidence from the methamphetamine literature [C and D below], given a lack of literature describing this phenomenon for illicit fentanyl). The resulting total range of bioavailability of smoked/vaporized fentanyl was 30.0% to 58.0%, represented by a uniform distribution ranging from 30.0% to 58.0%.

a. MacLeod, D.B., Habib, A.S., Ikeda, K., Spyker, D.A., Cassella, J.V., Ho, K.Y., Gan, T.J., 2012. Inhaled Fentanyl Aerosol in Healthy Volunteers: Pharmacokinetics and Pharmacodynamics. Anesthesia & Analgesia 115, 1071–1077. https://doi.org/10.1213/ANE.0b013e3182691898

i. This study estimated bioavailability of 96.8%, standard error of 7.62%.
b. Mather, L. E., Woodhouse, A., Ward, M. E., Farr, S. J., Rubsamen, R. A. & Eltherington, L. G. (1998). Pulmonary administration of aerosolised fentanyl: pharmacokinetic analysis of systemic delivery. *British Journal of Clinical Pharmacology, 46* (1), 37-43. i. Estimated bioavailability as 78%-100% depending on dose, using specialized device to vaporize fentanyl.
c. Cook CE, Jeffcoat AR, Hill JM, et al. Pharmacokinetics of methamphetamine self-administered to human subjects by smoking S-(+)-methamphetamine hydrochloride. Drug Metab Dispos. 1993;21(4):717-723.

i. The authors estimated 26.9% of methamphetamine was left in the pipe by users.
d. Harris DS, Boxenbaum H, Everhart ET, Sequeira G, Mendelson JE, Jones RT. The bioavailability of intranasal and smoked methamphetamine. Clin Pharmacol Ther. 2003;74(5):475-486. doi:10.1016/j.clpt.2003.08.002.

i. In this study, 45% of methamphetamine was estimated to be left in the pipe.
4. The bioavailability of intranasal (“snorted”) fentanyl was estimated using the below studies, which had estimates ranging from 55% to 89%. We additionally assumed that street grade drugs have an additional loss of efficiency due to impurities not present in the below studies using pharmaceutical grade drugs. We ultimately used a range of bioavailability values represented with a uniform distribution spanning 50% to 66%.

a. Nar di-Hiebl S, Ndieyira JW, Al Enzi Y, et al. Pharmacokinetic Characterisation and Comparison of Bioavailability of Intranasal Fentanyl, Transmucosal, and Intravenous Administration through a Three-Way Crossover Study in 24 Healthy Volunteers. *Pain Res Manag*. 2021;2021:2887773. doi:10.1155/2021/2887773

i. Mean estimate 74.70%, standard error of 3.46%.
b. Foster, D., Upton, R., Christrup, L., Popper, L., 2008. Pharmacokinetics and Pharmacodynamics of Intranasal Versus Intravenous Fentanyl in Patients with Pain after Oral Surgery. Ann Pharmacother 42, 1380–1387. https://doi.org/10.1345/aph.1L168

i. Estimate of 89.0%.
c. Striebel HW, Kramer J, Luhmann I, Rohierse-Hohler I, Rieger A. Pharmakokinetische Studie zur intranasalen Gabe von Fentanyl. Der Schrnerz 1993;7:122-5.

i. Range of 55%-71% depending on the pH of the formulation.
5. The bioavailability of injected fentanyl would be, by definition, 100%. However we added a correction factor for ‘missed’ shots that are not delivered intravenously, and instead are absorbed subcutaneously. We represented the bioavailability of injected fentanyl with a uniform distribution spanning 80% to 100%.
6. Among the n=47 individuals providing route of administration information alongside quantity of consumption information (resampled to create 1,000,000 draws) bioavailability was calculated for each draw. If an individual reported multiple routes of administration, then it was assumed that on average individuals evenly split their consumption between the different methods they reported. This could underestimate or overestimate their total absorption depending on their true route of administration habits and the relative frequencies of each method of consumption. Nevertheless the uncertainty in this fraction was incorporated into the model, by leveraging uniform distributions of each route of administration’s fraction of use. This did not change the point estimate of bioavailability versus simply taking an average, but it did increase the uncertainty in bioavailability (width of the distribution). This was accomplished by the following steps:

a. A weight was assigned to each route of administration, drawn from a uniform distribution spanning 0.2 to 0.8.
b. For each draw, the sum of all weights was calculated. Each weight was divided by this sum to rake the total weight to add to 1.0.
c. The final bioavailability for each draw was calculated as a weighted mean across the present routes of administration.
7. Milligrams of morphine equivalence factors between IV fentanyl and IV morphine were obtained from the below literature sources. Equivalence was defined as ranging from 33 to 100.

a. Ing, M.C., Keane, O.A., Lakshmanan, A., Kim, E., Lee, H.C., Kelley-Quon, L.I., 2024. Opioid equipotency conversions for hospitalized infants: a systematic review. J Perinatol 44, 1709–1718. https://doi.org/10.1038/s41372-024-02121-z.

i. This systematic review summarized n=8 studies from adult patients (excluding those describing pediatric populations), which had MME conversion factors between IV fentanyl and IV morphine ranging from 33 to 100, with n=6 studies reporting a factor of 100, n=2 reporting a factor of 50 to 100, and n=1 reporting a factor of 33.
8. The equivalence between oral morphine and IV morphine was obtained from the following sources, and was therefore defined as ranging from 2 to 3.

a. Mercadante, S., Villari, P., Ferrera, P., Casuccio, A., Fulfaro, F., 2002. Rapid titration with intravenous morphine for severe cancer pain and immediate oral conversion. Cancer 95, 203–208. https://doi.org/10.1002/cncr.10636.

i. The IV to PO equivalence was defined as ranging from 1:2 to 1:3.
b. Fortnightly Review: Morphine in cancer pain: modes of administration, 1996.. BMJ 312, 823. https://doi.org/10.1136/bmj.312.7034.823.

i. The IV to PO equivalence was defined as 1:3.
c. Starlander, J., Melin-Johansson, C., Jonsson, H., Axelsson, B., 2011. Oral-Parenteral Conversion Factor for Morphine in Palliative Cancer Care: A Prospective Randomized Crossover Pilot Study. Pain Research and Treatment 2011, 1–5. https://doi.org/10.1155/2011/504034.

i. The IV to PO equivalence was defined as 1:2, with a standard of 1:3 also recognized as reasonable.
9. MME factors converting from IV fentanyl to oral morphine were obtained by multiplying the conversion factors in steps 7 and 8 above, yielding a final distribution ranging from 66 to 300. This was represented by a uniform distribution, with a range of 66 to 300.

### Calculations

1. At the draw-level, correlation between quantity of fentanyl consumed, and route-of-administration, is preserved from the underlying data, as these quantities are sampled together.
2. For sensitivity analysis #1, inverse correlation is induced between quantity of fentanyl consumed and purity of fentanyl consumed, by sorting draws. This is accomplished by first sorting all 1,000,000 draws of all parameters by the quantity of fentanyl consumed (with routes of administration linked at the draw-level to maintain correlation from the underlying data). Subsequently, draws of fentanyl purity are removed, sorted in the inverse direction, and re-attached to the database. Supplemental Figure 1 below visualizes this process.
3. Calculations were performed according to the equation listed in the main text for the 1,000,000 draws of each parameter, derived as defined above. The distribution of quantity consumed, purity, and MME was graphed directly across the draws, and summary statistics (median and range) were calculated for each parameter.
4. For sensitivity analysis #1, the draws of fentanyl purity inversely correlated with quantity are used. All other parameters remain the same as the main analysis.
5. For sensitivity analysis #2, the original unsorted draws of fentanyl purity are used. All other parameters remain the same as the main analysis.
6. The primary estimates of MME are produced by combining the distributions created for sensitivity analyses #1 and #2, yielding 2,000,000 draws of MME. Summary statistics are calculated across all 2,000,000 draws.
7. For sensitivity analysis #3, bioavailability is set to the values corresponding to smoking for all individuals that reported smoking (regardless of other routes reported). As this represents 46/47 participants, this has the effect of essentially assuming that all participants only smoke fentanyl. All other model inputs remained the same as the main model.
8. For sensitivity analysis #4, a “noisy” inverse correlation is induced between the quantity of fentanyl consumed and fentanyl purity to simulate imperfect titration behavior. This is achieved by ranking both vectors in opposite directions, applying a uniform noise function X∼Uniform(-k/2,k/2) to the ranks of the purity distribution, and resorting the purity values by this noisy rank. K was defined as 0.9 * 1,000,000 draws. All other parameters remain the same as the main analysis.
9. For sensitivity analysis #5, a maximally conservative approach is used by combining the assumptions of sensitivity analysis #1 and sensitivity analysis #3. This model utilizes the perfectly inversely correlated draws of fentanyl purity alongside the smoking-only bioavailability values for all individuals who reported smoking. All other model inputs remain the same as the main model.

### Supplemental Results

**Supplemental Figure 1.**
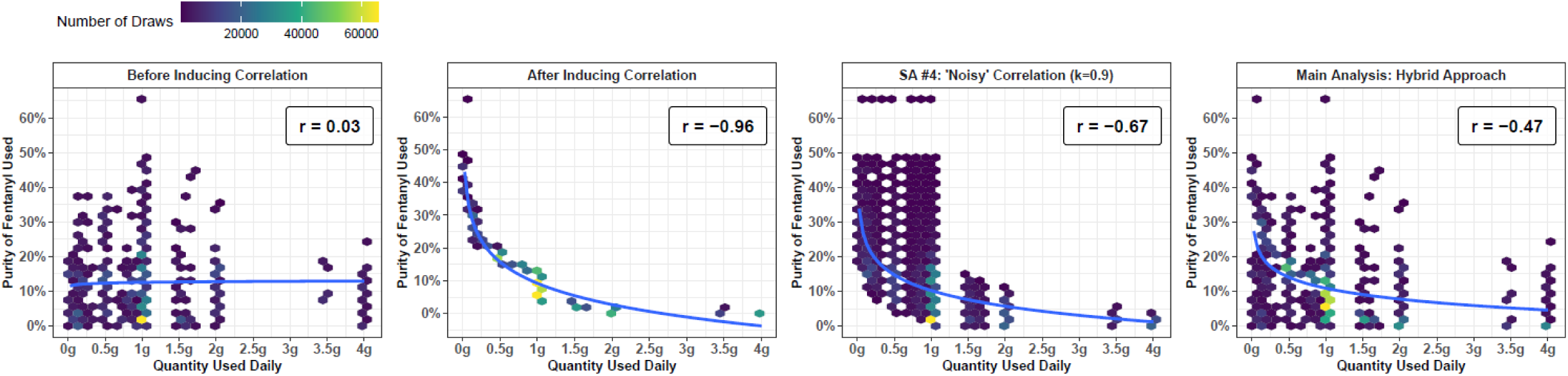
Purity and Quantity of Fentanyl Under Varying Correlation Assumptions. ?The modeled relationship between the purity and daily quantity of fentanyl consumed is shown across four panels, reflecting different methodological assumptions. Panel A displays the uncorrelated distribution before inducing correlation (used in Sensitivity Analysis #2). Panel B displays the distribution after inducing a maximal inverse correlation (used in Sensitivity Analysis #1). Panel C displays a noisy inverse correlation, representing imperfect titration behavior (used in Sensitivity Analysis #4). Panel D displays the hybrid approach used in the main analysis, which combines the draws from Panels A and B. Because these panels summarize large numbers of points (1,000,000 draws for Panels A–C; 2,000,000 draws for Panel D), hexbin plots are used instead of traditional scatter plots. The color of each bin indicates the number of draws summarized at that specific coordinate. In each panel, a log-linear line of best fit is shown using the formula purity ∼ log(quantity), and the corresponding Pearson correlation coefficient (r) is displayed in the top right corner. Correlation coefficients also reflect the log-linear relationship in each case.

**Supplemental Figure 2.**
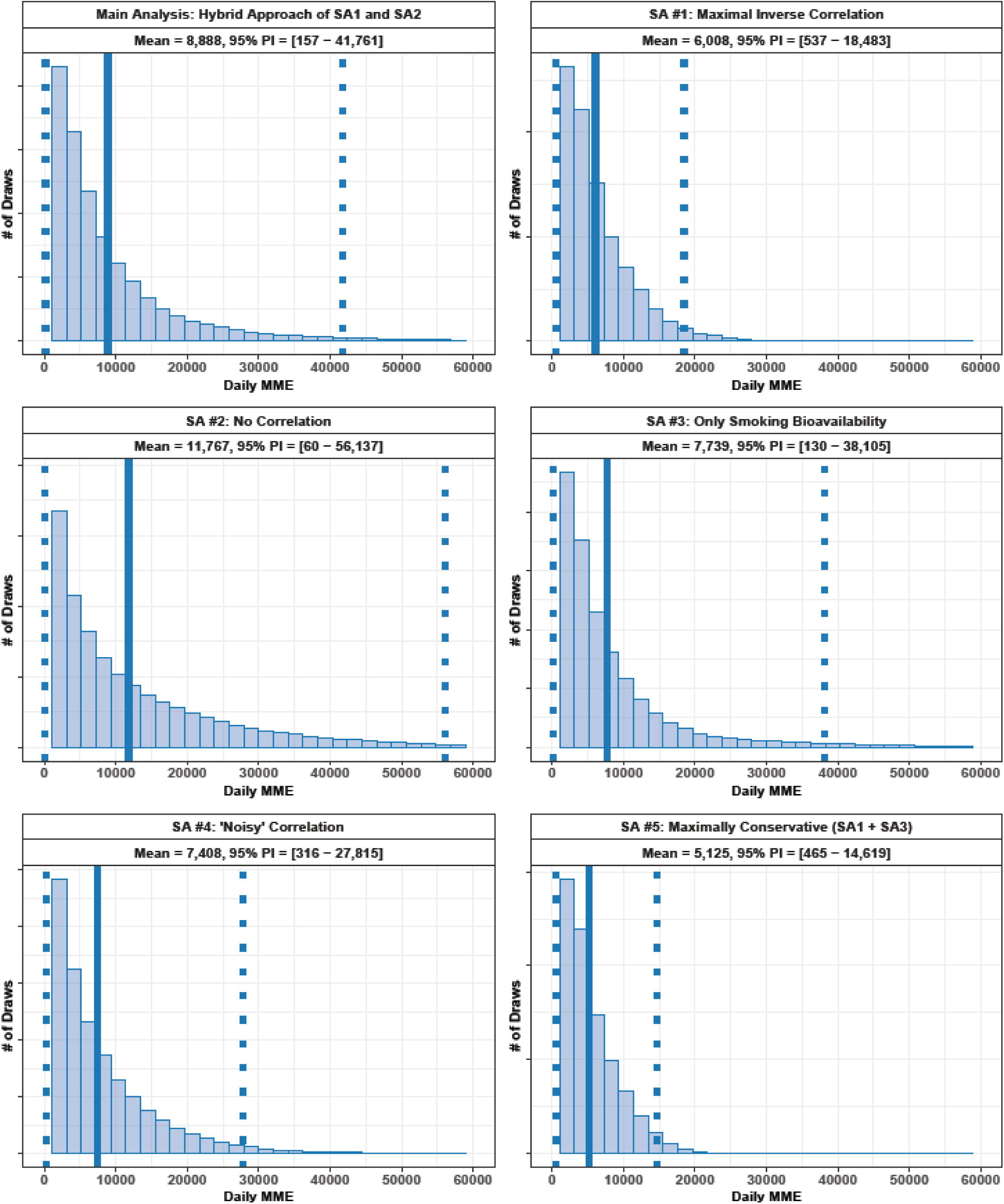
Estimated Daily MME Across Main and Sensitivity Analyses. The distribution of estimated daily fentanyl consumption in MME is shown for the main analysis and five sensitivity analyses. The panels display the model output for the main analysis (combining draws from SA1 and SA2); Sensitivity Analysis #1 (inducing maximal inverse correlation between quantity and purity); Sensitivity Analysis #2 (assuming no correlation); Sensitivity Analysis #3 (assuming bioavailability reflects only smoking); Sensitivity Analysis #4 (inducing a “noisy” inverse correlation); and Sensitivity Analysis #5 (a maximally conservative approach combining the assumptions of #1 and #3). In each panel, the distribution of the corresponding MME draws is shown as a histogram. A vertical solid line indicates the distribution mean, while dotted lines indicate the 95% prediction interval (2.5th and 97.5th percentiles). The calculated mean and 95% prediction interval values are annotated on a second title line within each panel.

**Supplemental Figure 3.**
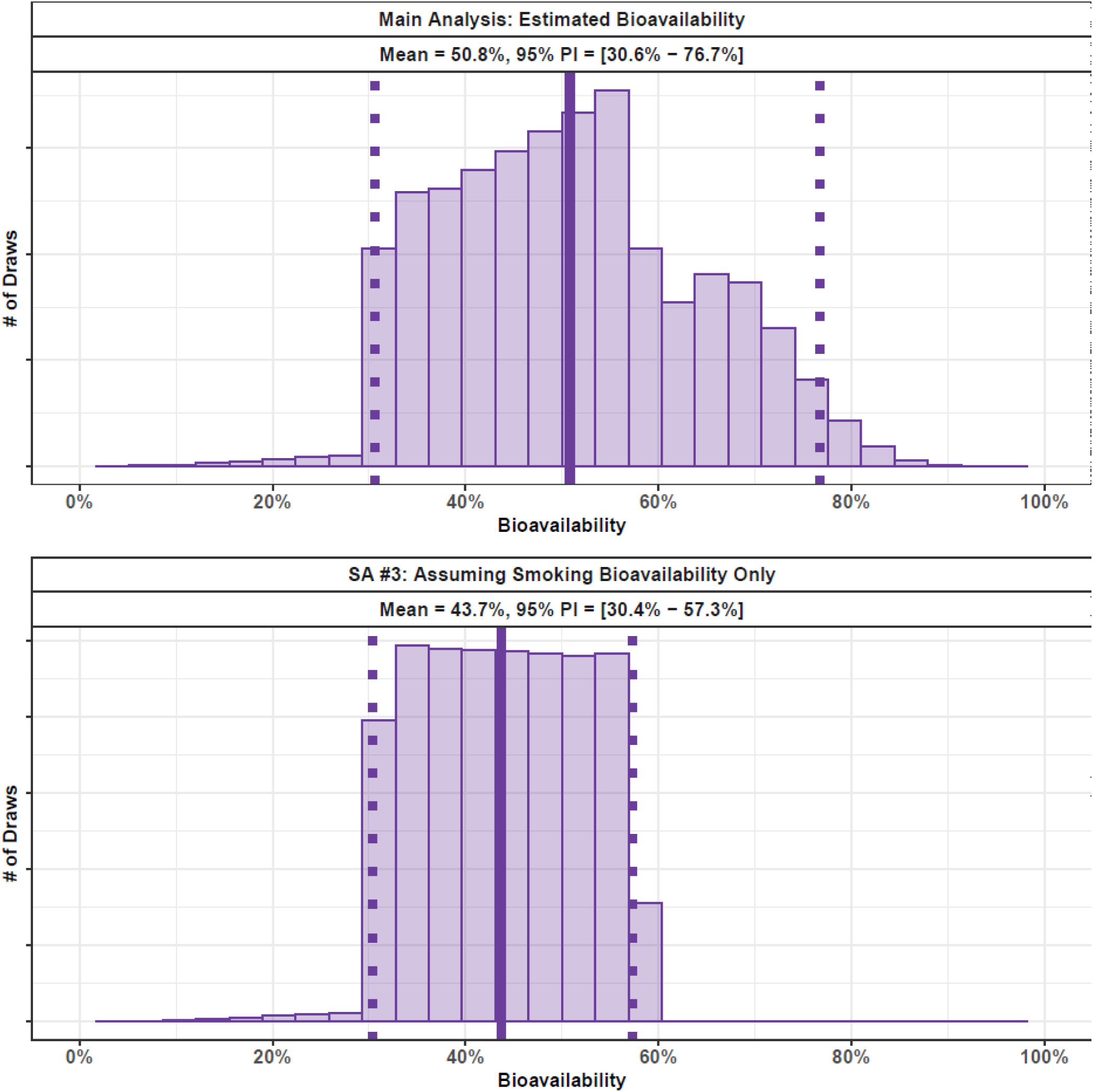
Estimated Bioavailability in Main Analysis vs. Sensitivity Analysis #3. The distribution of estimated bioavailability is compared between the main analysis (top) and the third sensitivity analysis (bottom). In the main analysis, bioavailability accounts for all reported routes of administration probabilistically. In Sensitivity Analysis #3, bioavailability values corresponding exclusively to smoking are applied to all individuals who reported smoking alongside other methods. In each panel, the distribution of the bioavailability draws is shown as a histogram. A vertical solid line indicates the distribution mean, while dotted lines indicate the 95% prediction interval (2.5th and 97.5th percentiles). Compared to the main analysis, the mean and overall width of the distribution in Sensitivity Analysis #3 are moderately reduced, reflecting the more constrained assumptions regarding absorption.

**Supplemental Figure 4.**
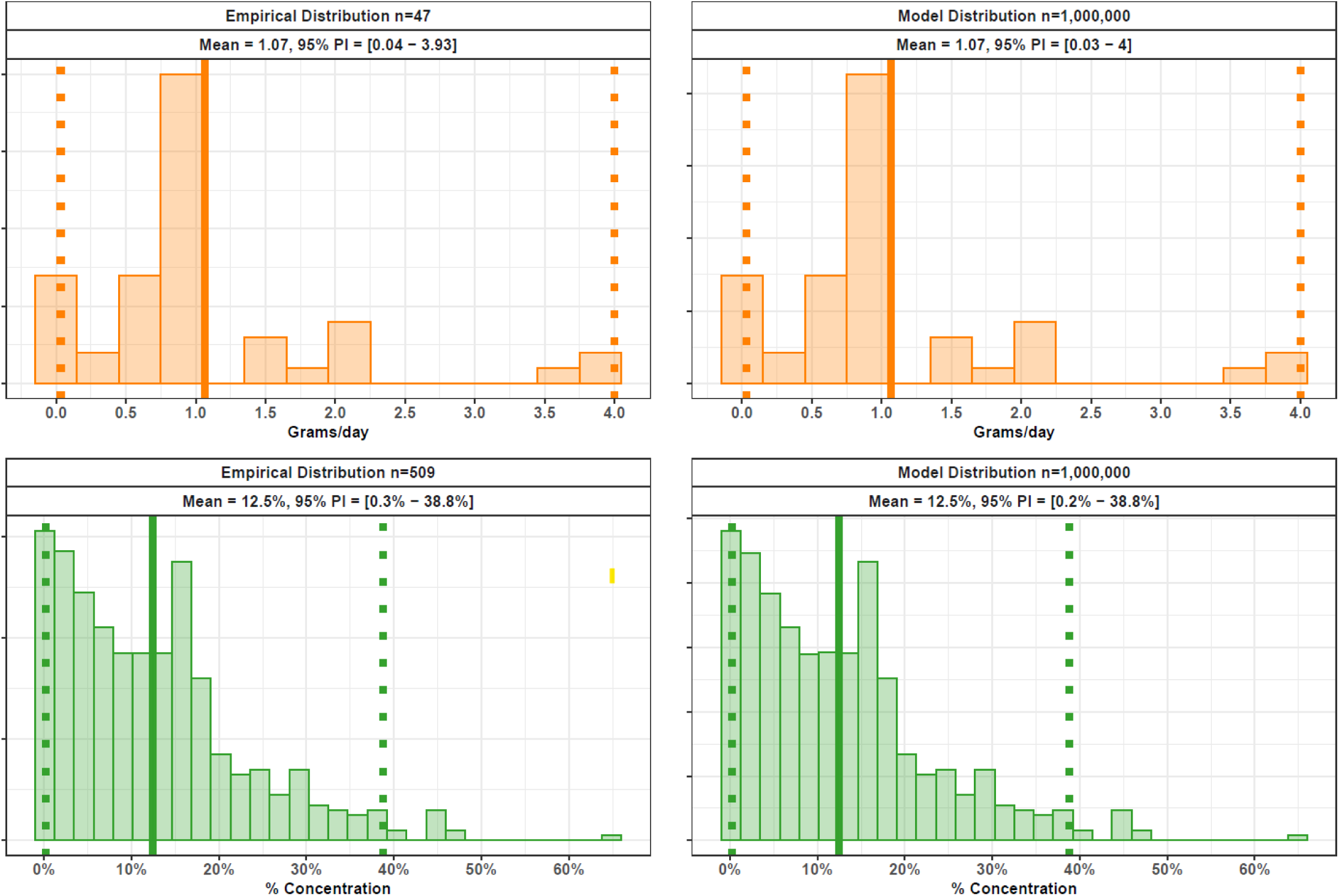
Empirical versus Model Distributions of Fentanyl Quantity and Purity. The empirical sample distributions are compared against the bootstrapped model distributions for daily fentanyl consumption and product purity. The top row displays the distribution of self-reported grams of illicit fentanyl consumed per day, comparing the raw empirical survey data (n=47, top left) to the resampled model draws (n=1,000,000, top right). The bottom row displays the percent concentration (purity) of expected fentanyl samples, comparing the empirical laboratory results (n=509, bottom left) to the resampled model draws (n=1,000,000, bottom right). In each panel, the distribution is shown as a histogram. A vertical solid line indicates the distribution mean, while dotted lines indicate the 95% prediction interval (2.5th and 97.5th percentiles). Due to the bootstrap resampling procedure with replacement, the empirical and model distributions are nearly identical.

## Notes

### Competing Interest Statement

The authors have declared no competing interest.

### Author Declarations

This project received approval by the UCLA IRB (IRB-22-0760).

### Summary of Updates

Added sensetivity analyses and supplemental figure.

